# Mutations on non-structural proteins of SARS-CoV-2 are possibly responsible for adverse clinical outcomes

**DOI:** 10.1101/2021.10.19.21265062

**Authors:** Takaya Ichikawa, Shiho Torii, Hikoyu Suzuki, Akio Takada, Satoshi Suzuki, Masahide Nakajima, Akihito Tampo, Yasutaka Kakinoki

## Abstract

Among a cluster of COVID-19 cases from the end of March through April 2021 in Asahikawa, we experienced the cases in which patients manifested severe clinical symptoms compared to patients who were infected before that. A hundred three patients (age range: 65 to 89 years old) enrolled in this study were divided into two groups, group A: the patients infected from November 2020 to March 2021, and group B: the patients in this cluster population. The mortality rates were 6.1% in group A and 16.2% in group B (OR: 2.97, 95%CI: 0.65-15.38). For the severity of disease, the patients in group B required higher oxygen flow rate in early course of admission (mild; p=0.892, moderate; p=0.117, severe; p=0.029). Whole viral genome sequences revealed five non-synonymous mutations by comparison of the isolates with each group. Of these, four were on non-structural proteins (NSPs) including nsp3, 6 and 15, and one was on S protein located near the C-terminus, suggesting that the mutations on NSPs could be responsible for adverse clinical outcomes in COVID-19 patients.

## Introduction

The Coronavirus disease 2019 (COVID-19), which is caused by Severe acute respiratory syndrome coronavirus 2 (SARS-CoV-2), initially broke out in Wuhan, China, on December 2019 and has subsequently spread around the world [1]. SARS-CoV-2 is an enveloped virus with a positive-sense single-stranded RNA genome of approximately 30 kb. Two-thirds of the viral genome at the 5’ terminus contains ORF1a and ORF1b, which encodes 16 non-structural proteins (NSPs). These NSPs play crucial roles in viral replication and evasion of host immune systems [2]. The other one-third of the 3’ end contains structural genes such as spike glycoprotein (S protein), envelope protein, membrane protein, nucleocapsid protein, and several accessory proteins [3]. Because replication of RNA viruses typically has a low fidelity, the viral genome of SARS-CoV-2 has accumulated mutations at an average of two nucleotides per month [4]. On 25th February 2021, World Health Organization (WHO) defined a variant of interest (VOI) as an isolate detected in several countries that changes phenotype under certain conditions, and a variant of concern (VOC) as an isolate in VOIs if it has been demonstrated to be associated with increase in transmissibility or virulence (https://www.who.int/emergencies/diseases/novel-coronavirus-2019). For example, VOC-202012/01 (Pango lineage B.1.1.7), as it is called “alpha variant”, has higher transmissibility and mortality compared to a reference isolate [5,6]. These variants, however, were not focused on mutations in NSPs, but in structural proteins, especially S protein. It remains unclear whether mutations in NSPs are responsible for the virulence of COVID-19.

As of October 2021, COVID-19 is responsible for 1.7 million cases and 18 thousand deaths in Japan (https://www.mhlw.go.jp/stf/covid-19/kokunainohasseijoukyou.html). The most remarkable clinical feature of this disease is its heterogeneity in clinical manifestations, ranging from no symptoms to critical illness [7]. Several risk factors for symptom severity have been known, such as age (over 65 years old), male sex, obesity, history of smoking, and comorbidities (including hypertension, diabetes mellitus, respiratory disease, cardiovascular disease, chronic kidney disease and cancer) [8,9,10,11]. In Japan, the several treatment options are currently recommended based on the symptom severity: Casirivimab and Imdevimab, remdesivir, dexamethasone and baricitinib (https://www.mhlw.go.jp/content/000815065.pdf). In clinical practice, methylprednisolone is also used for critically ill patients when they have a poor response for the dexamethasone therapy or when baricitinib is not available [12].

In Asahikawa city, Hokkaido prefecture located in northern part of Japan, A cluster of COVID-19 cases occurred in several bars where a group of customers enjoyed “karaoke” during daytime from the end of March through April 2021 (referred to bellow as cluster K), and 66 cases were involved into the infection according to the city’s survey (https://www.city.asahikawa.hokkaido.jp/kurashi/135/136/150/index.html). We found that the progression of disease among the patients in this cluster were faster compared to patients who were infected before that. Therefore, in order to identify factors associated with the progression of disease, we examined the viral genome sequences along with the clinical characteristics and treatment strategies, and then compared those with other cases.

## Material and Method

### Participants

All adult patients with COVID-19 who were admitted to Asahikawa City Hospital from November 2020 to April 2021 were enrolled in this retrospective study. All positive results were confirmed by real-time RT-PCR for the presence of SARS-CoV-2 in saliva or nasopharyngeal swab samples. The recovered patients were discharged from the hospital following the discharge criteria provided by Japanese Ministry of Health, Labour and Welfare when two criteria have met:10 days passed from the symptom onset and 72 hours passed from the symptom resolution, or when two consecutive negative PCR results are confirmed. Opt-out consent was obtained instead of written informed consent. We provided the patients with information explaining the proposed research plan via the website of Asahikawa City Hospital, and with the opportunity to opt out.

### Clinical data collection and definitions

The epidemiological data, medical history, underlying comorbidities, symptoms and signs at admission, oxygen flow rate, treatment and clinical outcomes were obtained from electronic medical records. Because the date of onset was unclear for some patients who were asymptomatic, we considered the date on which positive PCR result was obtained as day 0. Clinical outcomes were followed up until discharge or death. The primary outcome was mortality rate, and the secondary outcome was oxygen flow rate as a severity of the disease; mild: ≥1 L/min, moderate: ≥5 L/min and severe: ≥10 L/min.

### Viral genome sequence

The stored RNA extracts were used as clinical samples. Fist-strand cDNA was synthesized by using a PrimeScript IV 1^st^ strand cDNA Synthesis Mix (TaKaRa Bio) with random hexamer primers and extracted RNA, according to the manufacture’s protocols. The whole-genome sequencing of SARS-CoV-2 was carried out as reported previously [13]. Briefly, a total of 10 gene fragments were amplified with synthesized cDNA, specific primer sets for SARS-CoV-2 and PrimeSTAR GXL DNA polymerase (TaKaRa Bio). Then, the amplified products were directly sequenced in both directions by using the ABI PRISM 3130 Genetic Analyzer (Applied Biosystems) with specific primers. Our raw sequence data were converted to FASTQ format by in-house Python script with BioPython module [14], followed by trimming first 50 bps and the region after 900 bps by FASTX-Toolkit (http://hannonlab.cshl.edu/fastx_toolkit/). Trimmed sequence reads were mapped to the SARS-CoV-2 reference genome (accession number MN908947.3).

### Phylogenetic analysis

More than one million SARS-CoV-2 published genome sequences whose length were between 29,000 - 30,000 bps were downloaded from National Center for Biotechnology Information (NCBI) via Application Programming Interface (API) for the Entrez Programming Utilities (E-utilities) (https://www.ncbi.nlm.nih.gov/books/NBK25497/). Sequences including less than 29,000 definite bases, neither ‘N’ nor other mixed bases, were discarded.

All SARS-CoV-2 genome sequence data, including our assemblies, were mapped to the reference genome (accession number MN908947.3). Based on the SAM format mapping output, SNPs and short INDELs were detected and categorized to synonymous, non-synonymous or intergenic mutations by in-house Perl scripts, which simply picked up the differences from the reference sequence for each genome. For the phylogenetic analysis, a typical genome sequence of each variant, named alpha, beta, gamma, delta, epsilon, zeta, eta, theta, iota, kappa, and lambda by WHO, were chosen from the published data based on the following criteria; neither ‘N ‘nor other mixed bases were included in open reading frames (ORFs), number of substitution sites in ORFs were less than 100, all non-synonymous mutations which characterize each variant were included, and all other substitutes observed in more than 95% of the samples that satisfy the above conditions were also included. The non-synonymous mutations which characterize each variant are shown in Table S1, created based on the mutation prevalence information from outbreak.info (https://outbreak.info/). ORF10 was excluded from the analyses because one of our assemblies did not completely cover that region. Based on the alignment, suitable nucleotide substitution models for each partition were selected by ModelTest-NG [15], followed by maximum likelihood phylogenetic analyses using the selected substitution models by RAxML-NG [16]. The constructed phylogenetic tree was drawn and edited by MEGA X [17].

### Statistical analysis

We performed group comparisons using Fisher’s exact test for categorical variables, and independent group t-test for continuous variables. The categorical variables were expressed as frequencies (percentage), and the continuous variables as the median with interquartile range. Odds ratios (OR) with 95% confidence interval were calculated for the rates of mortality and the required oxygen flow rate. Cumulative probabilities of oxygen therapy were described with using the Kaplan-Meier analysis and compared using log-rank test. P-values of 0.05 or less were considered statistically significant. All statistical analyses were performed with EZR (Saitama Medical Center, Jichi Medical University, Saitama, Japan, version 1.54) [18], which is a graphical user interface for R software (The R Foundation for Statistical Computing, Vienna, Austria).

## Result

A total of 191 patients diagnosed with COVID-19 were admitted to our hospital from November 2020 to April 2021. The patients were divided into two groups, group A: the patients infected from November 2020 to March 2021, and group B: the patients whose infection was traced from the cluster K. We included the patients whose ages range between 65 and 89 years old for the following reasons; 1) patients who are younger than 65 years old are not necessarily requested for hospitalization in our city’s health care system, 2) in group A, many patients over 90 years old were transferred from a convalescent hospital where COVID-19 outbreak had occurred, thus the age profiles would be much different from group B if the patients aged over 90 years old were included in this group. Finally, 66 patients were assigned to group A and 37 patients to group B (Figure 1).

**Figure 1.**
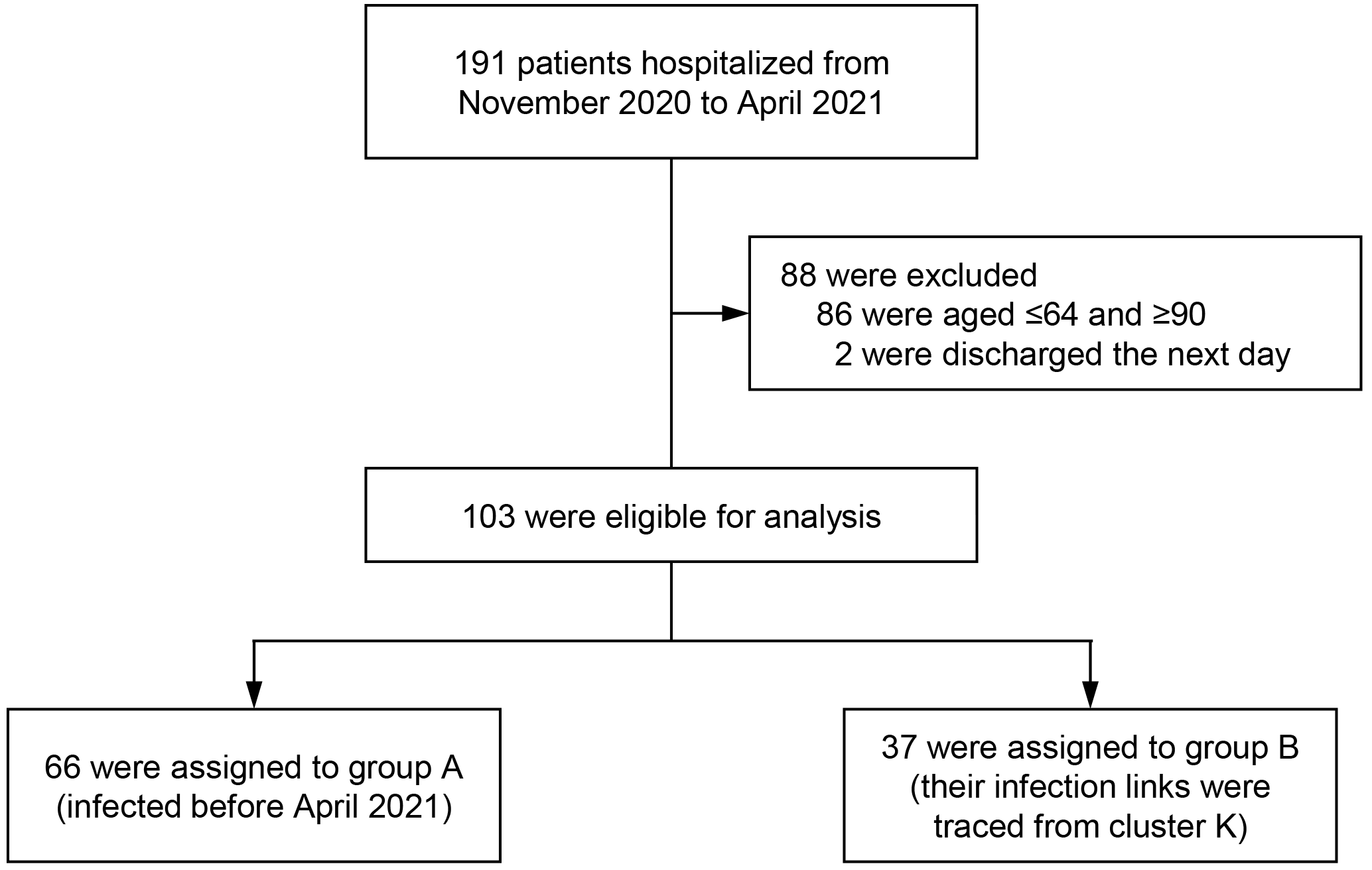
Flow diagram indicating the recruitment process in this study.

The clinical characteristics and treatment strategies used during hospitalization are shown in Table 1. Although there were no significant differences in clinical characteristics between the two groups, obesity and lifestyle-related diseases such as hypertension, dyslipidemia and diabetes tended to be more prevalent in group B. In treatment strategies, remdesivir tended to be used more frequently in group A and methylprednisolone in group B.

**Table 1.** Characteristics and treatment options of COVID-19 patients.

| Characteristic | Group A<br>(N=66) | Group B<br>(N=37) | P value |
| --- | --- | --- | --- |
| Age, median (IQR) | 76 (71-85) | 78 (74-82) | 0.75 |
| Male sex (%) | 32 (48.5) | 17 (45.9) | 0.84 |
| Duration of hospitalization,<br>median (IQR) | 12.5 (9-16) | 11 (59-16) | 0.85 |
| Obesity, BMI $\geq 30$ (%) | 4 (6.1) | 6 (16.2) | 0.16 |
| Current or ex-smoker (%) | 21 (31.8) | 13 (35.1) | 0.83 |
| Comorbidities (%) |  |  |  |
| Hypertention | 38 (57.6) | 28 (75.7) | 0.09 |
| Dyslipidemia | 21 (31.8) | 16 (43.2) | 0.29 |
| Diabetes mellitus | 12 (18.2) | 10 (27.0) | 0.32 |
| Chronic pulmonary disease | 12 (18.2) | 11 (29.7) | 0.22 |
| Cardiovascular disease | 10 (15.2) | 7 (18.9) | 0.78 |
| Chronic kidney disease | 10 (15.2) | 6 (16.2) | 1.00 |
| Cancer | 9 (13.6) | 2 (5.4) | 0.32 |
| Treatment options (%) |  |  |  |
| Dexamethasone | 39 (59.1) | 19 (51.4) | 0.54 |
| Methylprednisolone | 10 (15.2) | 11 (29.7) | 0.12 |
| Remdesivir | 8 (12.1) | 2 (5.4) | 0.32 |

IQR: interquartile range, BMI: body mass index

Four patients of 66 patients (6.1%) in group A and six of 37 patients (16.2%) in group B died from COVID-19 (OR: 2.97, 95%CI: 0.65-15.38) (Table 2). When limited to males, the differences in number of deaths became larger (6.3% vs 29.4%, OR: 5.99, 95%CI: 0.84-71.2). The clinical characteristics and the treatment of 10 deceased patients are shown in Table S2. Cumulative probability of the patients who required minimal oxygen therapy (mild) were similar in both groups (Table 2, Figure 2A). However, the patients in group B were likely to require more oxygen therapy in earlier course of hospitalization (Figure 2B, 2C). There was no significant difference in the cumulative probability between two groups for moderate oxygen therapy (p=0.117), but for severe oxygen therapy, the significant difference was observed (p=0.029). As with mortality, males in group B showed worse outcomes in the severity of disease (Table 2).

**Table 2.**
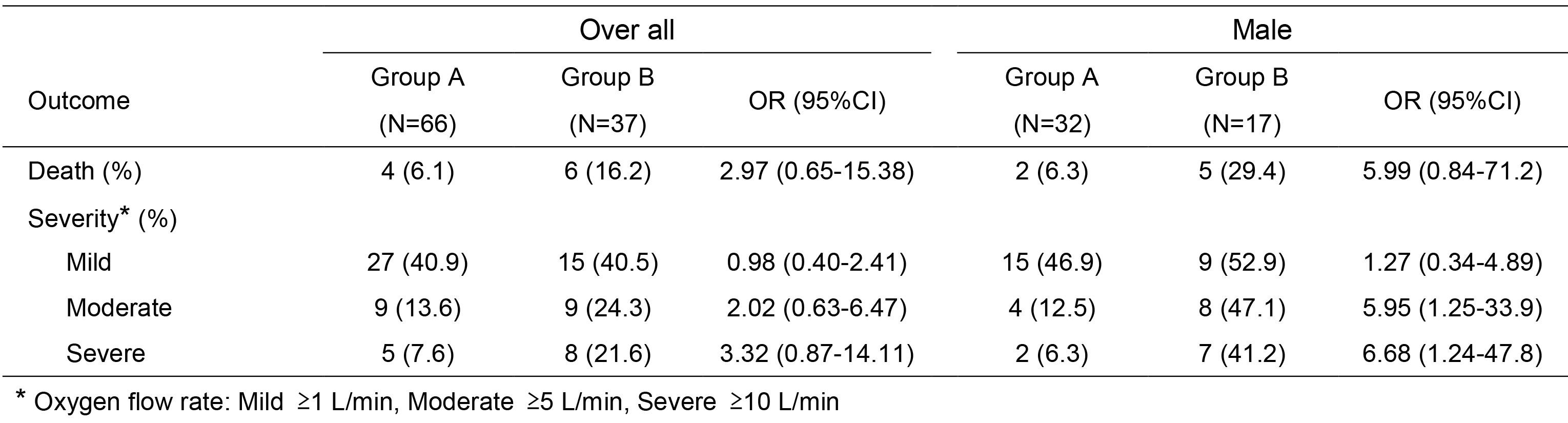
Primary and secondary outcomes of COVID-19 patients.

| Outcome | Over all |  |  | Male |  |  |
| --- | --- | --- | --- | --- | --- | --- |
|  | Group A<br>(N=66) | Group B<br>(N=37) | OR (95%CI) | Group A<br>(N=32) | Group B<br>(N=17) | OR (95%CI) |
| Death (%) | 4 (6.1) | 6 (16.2) | 2.97 (0.65-15.38) | 2 (6.3) | 5 (29.4) | 5.99 (0.84-71.2) |
| Severity* (%) |  |  |  |  |  |  |
| Mild | 27 (40.9) | 15 (40.5) | 0.98 (0.40-2.41) | 15 (46.9) | 9 (52.9) | 1.27 (0.34-4.89) |
| Moderate | 9 (13.6) | 9 (24.3) | 2.02 (0.63-6.47) | 4 (12.5) | 8 (47.1) | 5.95 (1.25-33.9) |
| Severe | 5 (7.6) | 8 (21.6) | 3.32 (0.87-14.11) | 2 (6.3) | 7 (41.2) | 6.68 (1.24-47.8) |

\* Oxygen flow rate: Mild $\geq 1$ L/min, Moderate $\geq 5$ L/min, Severe $\geq 10$ L/min

**Figure 2.**
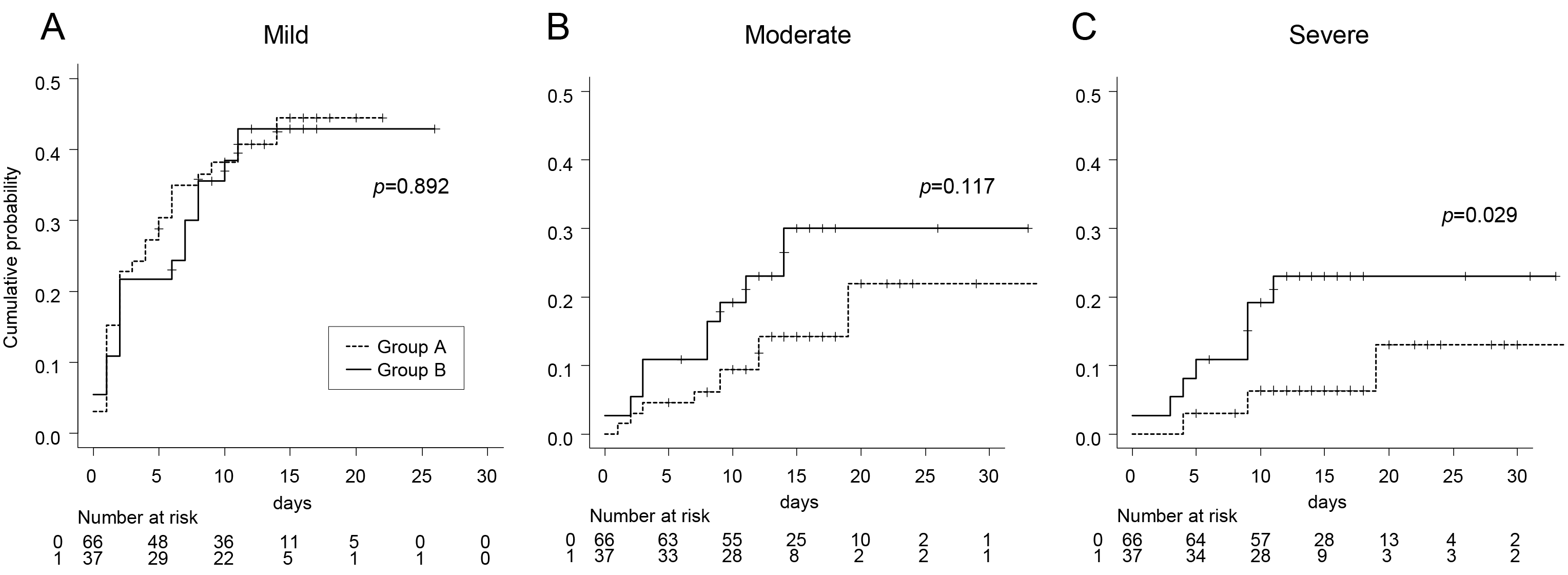
Cumulative probability of the patients who required oxygen therapy with mild: ≥1 L/min, moderate: ≥5 L/min and severe: ≥10 L/min. The log-rank test was used for statistical analysis.

Assuming that these differences in clinical outcomes would be due to viral factors, we examined the whole viral genome sequences representing two samples in each group, named Asahikawa_0108 and Asahikawa_0122 isolated in January 2021, Asahikawa_0404 and Asahikawa_0417 isolated in April 2021. The mutation sites which differed in these four samples are summarized in Table 3. Focusing on the non-synonymous mutations, five amino acid changes were identified (represented in bold). Of these, four amino acid changes were located on NSPs and one was on S protein, as illustrated in the viral genome scheme of SARS-CoV-2 (Figure 3). In order to assess strain relatedness, the phylogenetic analysis was carried out. The isolates in this study were closely related to the strain previously isolated in Japan (accession number BS000756.1; Pango lineage B.1.1.214), but not to any of the variants labeled by WHO (Figure 4).

**Table 3.** Mutation sites that differed in the four samples.

| Nucleotide position | Gene | Group A |  | Group B |  | Amino acid change |
| --- | --- | --- | --- | --- | --- | --- |
|  |  | Asahikawa_0108 | Asahikawa_0122 | Asahikawa_0404 | Asahikawa_0417 |  |
| 4233 | Orf1a | G | A | A | A | D1323G |
| 6380 |  | C | T | C | C | L2039F |
| 6433 |  | T | C | C | C | Syn |
| 6532 |  | G | G | T | T | **E2089D** |
| 6997 |  | T | C | C | C | Syn |
| 8371 |  | T | G | G | G | Q2702H |
| 8917 |  | C | T | T | T | Syn |
| 9207 |  | T | C | C | C | S2981F |
| 10042 |  | A | A | G | G | Syn |
| 11058 |  | C | C | T | T | **T3598I** |
| 11430 |  | A | A | G | G | **T3722C** |
| 12049 |  | C | T | T | T | Syn |
| 13366 |  | C | C | T | T | Syn |
| 15240 | Orf1b | T | C | C | C | Syn |
| 15957 |  | G | A | G | G | Syn |
| 17502 |  | C | T | C | C | Syn |
| 19894 |  | G | G | T | T | **A2143S** |
| 20658 |  | A | G | A | A | Syn |
| 22502 | S | C | A | C | C | Q314K |
| 23191 |  | C | C | T | T | Syn |
| 23587 |  | G | T | T | T | Q675H |
| 23720 |  | G | A | A | A | I720V |
| 24748 |  | C | C | T | T | Syn |
| 25020 |  | A | A | G | G | **D1153G** |
| 29353 | N | C | T | T | T | Syn |

Position number based on alignment to NCBI reference sequence: NC\_045512 (Wuhan-Hu-1).
The different parts between the two samples in group A are shown in a gray area. Amino acid changes identified between two groups were represented in bold.

**Figure 3.**
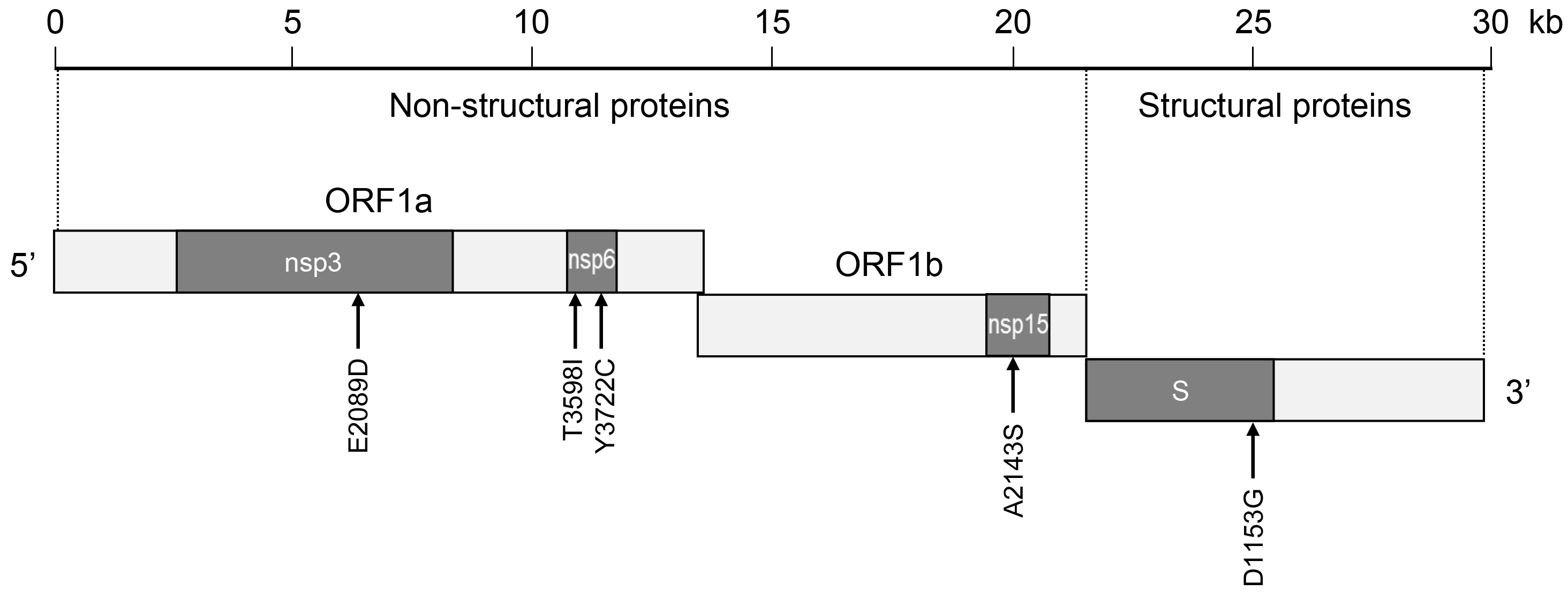
Scheme of SARS-CoV-2 genome. The non-synonymous mutation sites are indicated by arrows.

**Figure 4.**
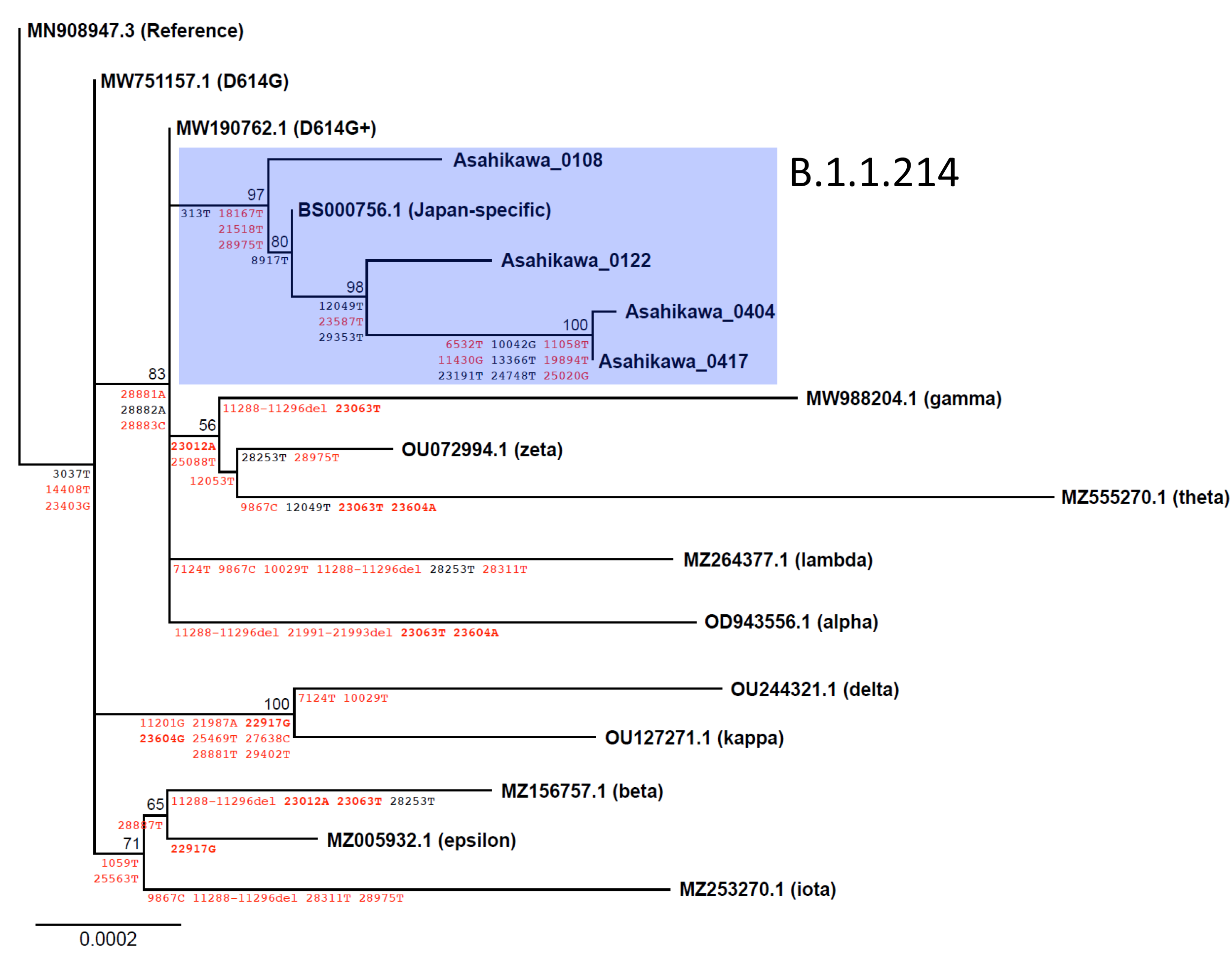
Maximum likelihood phylogenetic tree of SARS-CoV-2 variants. Black and rend characters indicate synonymous and non-synonymous mutations respectively. Red bold characters indicate mutations of interest. Numbers on internal nodes are bootstrap values for each divergence.

## Discussion

We here experienced the cases of a single COVID-19 cluster in which the conventional treatment was unsuccessful and the hospitalized patients became more severely ill, thus performed the retrospective analysis to find out the causative factors. The patients in group B had a higher mortality rate compared to group A, but no significant difference was indicated. For the severity of disease, the patients in group B required higher oxygen flow rate in early course of admission. Furthermore, the differences became larger when limited to males, with 41.2% of male severe patients in group B against 6.3% in group A.

In terms of treatment strategies, we were likely to use methylprednisolone monotherapy for severe cases in group B. This was because we had experienced that, in the winter of 2020, patients who received methylprednisolone therapy appeared to have a better response than those who were treated with remdesivir after poor response for dexamethasone, although the treatment guideline for COVID-19 in Japan recommends the use of remdesivir for patients with pneumonia whose oxygen saturation falls below 96% (https://www.mhlw.go.jp/content/000815065.pdf). In our study, there was no significant difference of remdesivir usage between two groups, thus the viral factors could have an impact on the clinical outcomes.

Whole viral genome sequences revealed five non-synonymous mutations by comparison of the isolates with each group. Interestingly, four of them were on NSPs. The sole structural mutation, D1153G on S protein, was located near the C-terminus and outside the range of three-dimensional structures obtained by crystallography, suggesting neither the head nor stalk of S protein. Therefore, D1153G is not expected to involve in the virulence of the disease. Of the four mutation sites on NSPs, two were found on nsp6 (T3598I and Y3722C), and the rest of them were on nsp3 (E2089D) and nsp15 (A2143S). To our knowledge, none of these mutations are known to be the factors that determine the severity of disease in clinical practice, and it is unclear whether the mutations affect the functions of these NSPs. However, all of them have been reported to play roles in suppression of type 1 interferon in host cells [19,20,21], and also nps6 involves in the formation of autophagosome in host cells [22]. The potential sites under positive selective pressure have been found on nsp6 near T3598I according to an evolutionary analysis on SARS-CoV-2 genome sequences of 351 clinical samples [23]. Therefore, the mutations on nsp3, 6 and 15 could be responsible for its function to interact with host immunity and autophagy. Further investigations are required to reveal their functions and involvement in the virulence.

The phylogenetic analysis revealed that the isolates in this study are derived from Pango lineage B.1.1.214 which was the dominant strain in so-called “the third wave”, the epidemic period from October to December 2020 in Japan [24]. Considering the fact that any strains identical to the viral sequence of group B cannot be found in more than 1,000,000 registered sequences in NCBI, it is likely that the virus evolved locally from January to April 2021 in Hokkaido. Since May 2021, alpha variant has been dominant throughout Japan including Hokkaido; 78% of the samples isolated from April 26 to May 2 2021 in Hokkaido were already alpha variant (https://www.mhlw.go.jp/content/10900000/000779013.pdf). As a consequence, B.1.1.214 strains in Asahikawa seems to be eliminated. This is similar to the situation in influenza A virus, for example, where pandemic influenza A (H1N1) pdm09 replaced Russian flu (H1N1) since 2009 [25,26].

There are some limitations to this retrospective study. First, the recommended therapy for COVID-19 have been changing dynamically. Because the best treatment strategies are selected based on the evidence at that time, it is difficult to compare clinical outcomes between different periods of time in real-life practice. Second, this is a single institution study; not all cases in cluster K have been assessed. The mortality and severity rate would be different when including the cases admitted to other hospitals. Third, we examined only two samples from each group in this study. Due to the limited human and financial resources, we were unbale to perform the viral genome analysis of much more samples.

In conclusion, the present study demonstrated that some viral factors, four non-synonymous mutations on nsp3, 6 and 15 but not on S protein, could be responsible for adverse clinical outcomes in COVID-19 patients. Though scientists and physicians all over the world have focused on the S protein mutations, in order to comprehensively understand the epidemiology of COVID-19, we should have an insight into other type of mutations such as those on NSPs.

## Supporting information

Spplemental tables

## Data Availability

All data produced in the present study are available upon reasonable request to the authors.

https://www.ncbi.nlm.nih.gov/books/NBK25497/

## Acknowledgment

We thank Prof. Yoshiharu Matsuura, Osaka University, for funding the viral sequence analysis. We also thank our medical team worked on COVID-19 and all of the staff at Asahikawa Health Center for their support.

