## Supplementary material for "Mutations on non-structural proteins of SARS-CoV-2 are possibly responsible for adverse clinical outcomes": Spplemental tables

Table S1. Non-synonymous mutations for characterizing the variants.

| | ORF1a | ORF1b | S | ORF3a | M | ORF7a | ORF8 | N |
|---|---|---|---|---|---|---|---|---|
| Alpha | T1001I<br>A1708D<br>I2230T | P314L | N501Y T716I<br>A570D S982A<br>D614G D1118H<br>P681H | | | | Q27*<br>R52I<br>Y73C | (D3L)<br>R203K<br>G204R<br>S235F |
| Beta | T265I<br>K3353R | P314L | E484K<br>N501Y<br>D614G | Q57H | | | | T205I |
| Gamma | S1188L<br>K1795Q | P314L<br>E1264D | L18F N501Y<br>T20N D614G<br>P26S H655Y<br>D138Y T1027I<br>E484K V1176F | S253P | | | E92K | P80R<br>R203K<br>G204R |
| Delta | P2287S<br>T3255I<br>T3646A | P314L<br>G662S<br>P1000L | T19R D614G<br>L452R P681R<br>T478K D950N | S26L | I82T | T120I | | D63G<br>R203M<br>D377Y |
| Epsilon | T265I | P314L | L452R<br>D614G | Q57H | | | | T205I |
| Zeta | L3468V<br>L3930F | P314L | E484K<br>D614G<br>V1176F | | | | | A119S<br>R203K<br>G204R<br>M234I |
| Eta | T2007I | (P314F) | A67V Q677H<br>E484K F888L<br>D614G | | I82T | | | T205I |
| Theta | L3201P<br>D3681E<br>L3930F | P314L<br>A1291V | E484K E1092K<br>N501Y H1101Y<br>D614G V1176F<br>P681H | | | | | R203K<br>G204R |
| Iota | T265I<br>L3201P | P314L | D614G | P42L<br>Q57H | | | T11I | M234I |
| Kappa | T1567I<br>T3646A | P314L<br>M1352I | L452R<br>D614G<br>P681R | S26L | | | | R203M<br>D377Y |
| Lambda | T1246I<br>P2287S<br>F2387V<br>L3201P<br>T3255I | P314L | T76I<br>L452Q<br>F490S<br>D614G<br>T859N | | | | | P13L<br>R203K<br>G204R<br>G214C |
| D614G | | P314L | D614G | | | | | |
| D614G+ | | P314L | D614G | | | | | R203K<br>G204R |
| Japan | | P314L<br>P1567L<br>R2684I | D614G | | | | | R203K<br>G204R<br>M234I |

Table S2. Characteristics and treatment of deceased patients

| Patient No. | Sex | Age | Duration of hospitalization | Obesity | Current or ex-Smoker | Comorbidity | Treatment |
|---|---|---|---|---|---|---|---|
| Group A | | | | | | | |
| 1 | Male | 70s | 3 | No | Yes | COPD, Cancer | DEX, RDV |
| 2 | Female | 70s | 19 | No | No | HTN | DEX, mPSL |
| 3 | Female | 80s | 28 | No | No | HTN, CVD | DEX, mPSL, RDV |
| 4 | Male | 80s | 6 | No | No | HTN, COPD, Cancer | DEX |
| Group B | | | | | | | |
| 1 | Male | 70s | 21 | No | No | HTN, COPD, CKD | mPSL,RDV |
| 2 | Female | 70s | 10 | Yes | Yes | HTN, DLP, COPD, CKD | mPSL |
| 3 | Male | 70s | 26 | Yes | No | HTN, DLP, DM | DEX, mPSL |
| 4 | Male | 70s | 21 | No | Yes | HTN, DM, CVD | DEX, mPSL |
| 5 | Male | 80s | 5 | No | Yes | DM, COPD | mPSL |
| 6 | Male | 80s | 16 | No | Yes | HTN, DM, CVD, CKD, Cancer | DEX, mPSL |

COPD: chronic obstructive pulmonary disease, HTN: hypertension, CVD: cardiovascular disease, DLP: dyslipidemia, DM: diabetes mellitus,
DEX: dexamethasone, RDV: remdesivir, mPSL: methylprednisolone
